# Utilization, satisfaction and barriers to access of health services for Arabic-speaking refugees resettled in Connecticut, USA after completion of Refugee Medical Assistance (RMA)

**DOI:** 10.1101/2021.07.05.21259732

**Authors:** Ali Elreichouni, Sarah Aly, Kaitlin Maciejewski, Islam Salem, Noah Ghossein, M. Salah Mankash, James Dziura, Hani Mowafi

**Author notes:** **CORRESPONDING AUTHOR:** Hani Mowafi, 464 Congress Ave, Suite 260, New Haven, CT 06519.

## Abstract

**Background:** Arabic-speaking refugees are the largest group of resettled persons in the United States since 2008, yet little is known about their rates of healthcare access, utilization and satisfaction.

**Methods:** From January to December 2019, a household survey was conducted of Arabic-speaking refugees resettled in CT. Data collected included demographics, prevalence of chronic conditions, patterns of health seeking behavior, insurance status and patient satisfaction using the Patient Satisfaction Questionnaire (PSQ-18).

**Results:** Sixty-five households responded to the survey representing 295 Arabic-speaking refugees – of which 141 (48%) were children. Forty-seven households (72%) reported members with chronic conditions, 62 persons (21%) needed daily medication, 285 (97%) persons were insured. Respondents reported high median patient satisfaction with wide variation.

**Conclusion:** Arabic-speaking refugees resettled in CT are young but have similar adult rates of chronic medical conditions as the host community. They majority remained insured after RMA lapsed. They expressed median high satisfaction with health services but with wide variation. Inaccessibility of health services in Arabic and difficulty obtaining medications remain areas in need of improvement.

## INTRODUCTION

The United States has resettled approximately three million refugees since 1975, the demographics of which vary with global events and trends[1]. As a result of recent conflicts in the Middle East and North Africa (MENA) Arabic-speaking refugees comprised the largest group resettled in the United States between 2008 and 2017[1]. According to the United Nations High Commissioner for Refugees (UNHCR), resettlement states are responsible for providing both legal and physical protection for refugees, along with political, social, and economic rights equivalent to those held by nationals. In reality, many refugees struggle to overcome cultural and linguistic barriers within their new homes, especially as they attempt to access healthcare services[2]. Surveys of refugees resettled with the United States have identified language and communication as common concerns among those attempting to access health services.[3-6] Only 7% of refugees report “good” English proficiency during pre-arrival screenings[7], and similar concerns – along with a sense of social disconnection – are prevalent among Arab refugees and immigrants[8]. Furthermore, when compared to US-born Arabs, Arab immigrants are more likely to self-report fair or poor levels of health – especially non-English speakers[9].

### Conceptual Framework

There is insufficient research on the physical and mental health needs of Arab-Americans overall, despite large communities in the US for well over a century[10-12]. Complicating study of this community’s health is the lack of an ethnicity identifier in standardized public surveys, vital statistics, and most electronic medical records – with Arab or Middle Eastern ethnicity being subsumed under White/Caucasian – which inhibits the use of large datasets to detect disparities in outcomes and access that can affect community health. Furthermore, recent events including global wars, terrorist attacks, and the political climate in the United States has led to a rise in Islamophobia and stigmatization of not only Muslims but of people suspected because of language or appearance to be from Muslim countries[6, 10, 13, 14]. Arabic is the primary language by the largest number of refugees resettled in the United States in the last dozen years. While Arabic language is spoken in a wide range of countries that exhibit ethnic and socio-economic diversity within and between them, we propose to use Arabic-speaking designation as a surrogate marker to identify this large group of resettled persons. Similar studies of physical and mental health have used this approach to identify this target population and to assess this community[14, 15]. Further, language and cultural barriers are routinely identified as obstacles to accessing care and overcoming them have been demonstrated to improve health access for Arabic-speaking refugees in discrete vertical health programs such as breast-cancer screening[13, 15].

The Refugee Medical Assistance program (RMA) provides short-term medical coverage for refugees and other persons who are eligible for Office of Refugees and Resettlement (ORR) benefits for eight months starting the date of arrival in the country or granting of asylum[16]. This study seeks to use household interviews with Arabic-speaking refugees resettled in Connecticut (CT) to identify and characterize barriers to their accessing health services; patterns of health-seeking behavior; and acceptability of health services in the period after Refugee Medical Assistance (RMA) has ended.

## METHODS

### Participants

From January 1^st^ – December 31^st^, 2019 a household survey was conducted of Arabic speaking refugees resettled in CT between January 2016 and June 2018. A contact list of 117 households representing 362 resettled persons was obtained from Integrated Refugee and Immigrant Services (IRIS) – the leading refugee resettlement agency in the State of Connecticut.

### Data Collection

A team of 5 trained research assistants contacted each household up to five times before listing them as “unable to reach”. All known phone numbers for the household and the phone number of any known US contact were contacted. All research assistants were medical trainees and were proficient in Arabic language. Upon contact with a household member, verbal consent was in Arabic or English at the discretion of the respondent. Then, an appointment was made to conduct the survey either in person or over the phone at the discretion of the respondent. If phone survey was preferred, survey questions were also provided in writing in both Arabic and English to the respondent prior to the appointment.

### Measures

The survey instrument was comprised of two parts. Part 1 included household demographics, insurance status of household members, information regarding chronic medical conditions and chronic medication use and patterns of recent health-seeking behavior. Part 2 assessed respondent satisfaction with health services using the Patient Satisfaction Questionnaire-18 (PSQ-18) (RAND Corporation, Santa Monica, CA)[17] (Appendix – Survey Questionnaire). All responses were entered in a deidentified dataset using Qualtrics survey software (Qualtrics, Provo, UT). Households that completed the survey were given a VISA gift card worth $50 in consideration of their participation and to offset the cost of their time and transportation to the interview.

### Analysis

Survey responses were analyzed using SPSS (IBM SPSS Statistics for Windows, Version 25.0, Armonk, NY). This study was approved by the Yale University Human Subjects Research Committee A HIPAA Waiver for written authorization was approved to allow authorization to be obtained verbally.

## RESULTS

Of 117 households listing Arabic as their primary language resettled in CT between January 2016 and July 2018, 5 were noted to have left the country, 22 had no available contact information, leaving 90 households who could be contacted for interview. Of these, 65 (72%) completed the survey corresponding to 295 individuals, 14 (16%) declined to participate, and 11 (12%) could not be contacted despite 5 separate attempts (Figure 1 – Household Recruitment).

**Figure 1:**
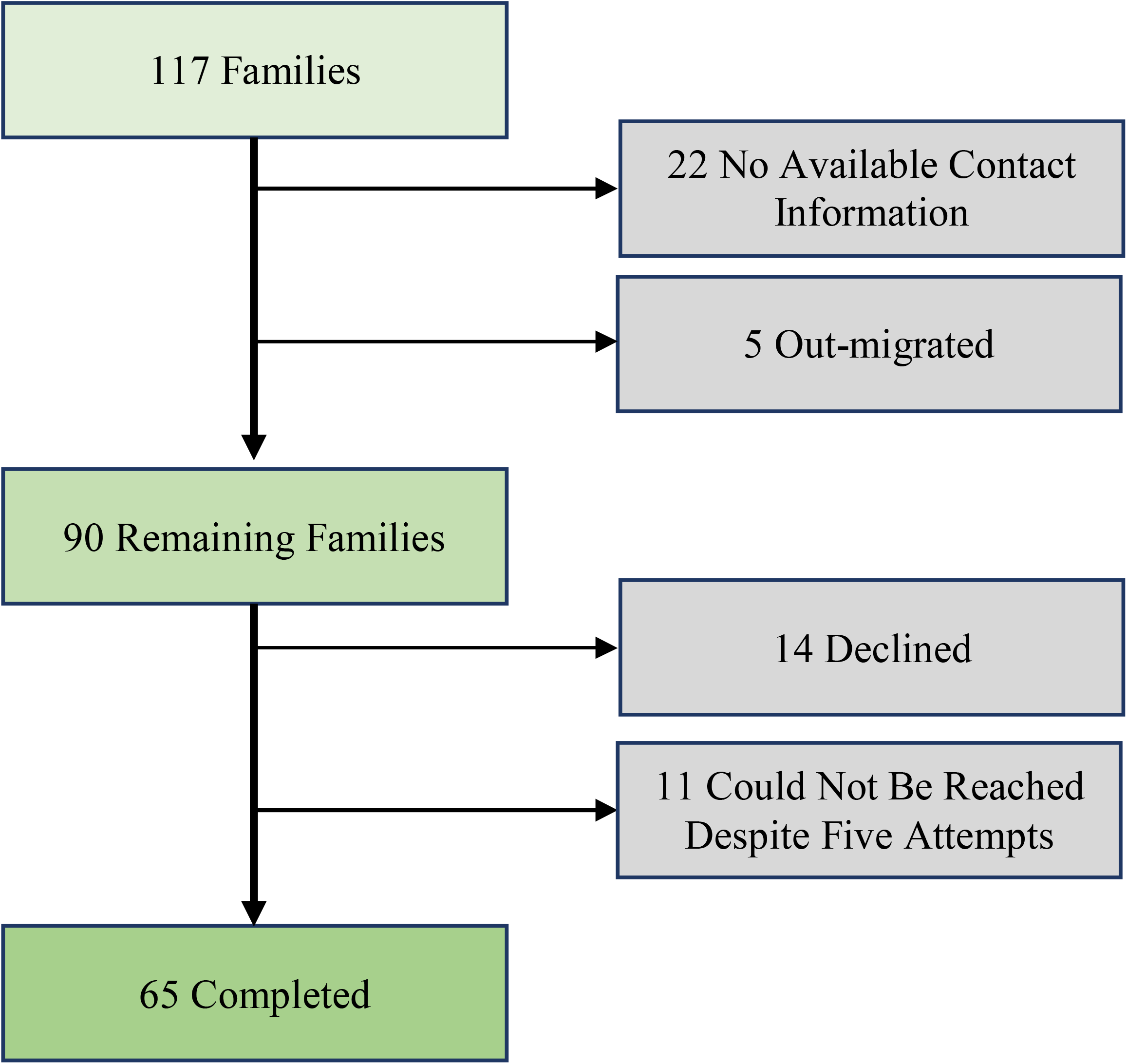
Survey Participant Composition.

Respondent household members were young with 141 (48%) children under 18 years of age, 148 (50%) adults and 6 (2%) elderly over 65 years of age (Table 1). Despite the predominance of young persons in surveyed households, 47 (72%) of households reported at least one family member with a chronic medical condition. Of 295 individuals represented, respondents reported 19 (6%) with high blood pressure, 17 (6%) with diabetes, 10 (3%) with cardiac conditions, 21 (7%) with chronic respiratory conditions, 19 (6%) with high cholesterol, 16 (5%) with chronic headaches, 7 (2%) with blood-related disorders, 1 (0.3%) with cancer, and 32 (11%) persons who reported other chronic conditions (Table 2). In addition, 25 household members (8%) reported functional limitation characterized as difficulty walking or needed assistance with activities of daily living (ADLs).

**Table 1:** Demographics and Characteristics of Arabic-Speaking Refugees.

|  | **N** | **%** |
| --- | --- | --- |
| **Females** | 177 | 60% |
| **Age (years)** |  |  |
| <18 | 141 | 48% |
| 18-24 | 38 | 13% |
| 25-34 | 38 | 13% |
| 35-55 | 41 | 14% |
| 45-54 | 20 | 7% |
| 55-64 | 11 | 4% |
| 65 and older | 6 | 2% |
| **Arrival Year** |  |  |
| 2016 | 47 | 72% |
| 2017 | 8 | 12% |
| 2018 | 10 | 15% |
| **Median Number of Adults (Range)** | 2 | (1 – 6) |
| **Median Number of Children (Range)** | 2 | (0 – 7) |
| **Health Insurance (Persons)** | 285 | 97% |

**Table 2:** Household chronic health profile.

|  | N | % |  |
| --- | --- | --- | --- |
| **Insurance Coverage(s) (No. Households)** |  |  |  |
| Medicaid/Husky Alone | 55 | 84.6% |  |
| Employer-Based Plan Alone | 1 | 1.5% |  |
| Medicaid/Husky & Private Insurance | 2 | 3.1% |  |
| Medicaid & Employer-based plan | 5 | 7.7% |  |
| No Insurance | 2 | 3.1% |  |
| **Primary Care-seeking venue (No. Households)** |  |  |  |
| Primary doctor | 53 | 81.5% |  |
| Walk in/ Urgent care | 6 | 9.2% |  |
| Emergency department | 5 | 7.7% |  |
| Other | 1 | 1.5% |  |
| **Median Health Visits per Household (prior 6 mo.)** |  |  | **Range** |
| Primary Care | 4 |  | (0 – 100) |
| Walk-in Clinic/Urgent Care | 0 |  | (0 – 8) |
| Emergency Department | 1 |  | (0 – 10) |
| Other | 0 |  | (0 – 13) |
| **HH reporting chronic condition** | 47 | 72.3% |  |
| **Persons Reporting Chronic Conditions** |  |  | *USA Prevalence\** |
| Hypertension | 19 | 6.4% | 29.9% |
| Diabetes | 17 | 5.8% | 9.8% |
| Heart Disease | 10 | 3.4% | 12.1% |
| Lung Disease | 21 | 7.1% | 6.2%, 9.5%<sup>+</sup> |
| High Cholesterol | 19 | 6.4% | 11.8% |
| Chronic Headaches | 16 | 5.4% | 15.3%<sup>§</sup> |
| Hematologic disorders | 7 | 2.4% | 5.6%<sup>▼</sup> |
| Cancer | 1 | 0.3% | 9.4% |
| Other Chronic Conditions | 32 | 10.8% | -- |
| **Difficulty with ADLs (Persons)** | 25 | 8.5% |  |
| **Requiring Daily Medications (Persons)** | 62 | 7.1% |  |
| **Difficulty Obtaining Daily Medications** | 17 | 5.8% |  |
| **Medication Payment (No. Households)** |  |  |  |
| Insurance | 51 | 78% |  |
| Insurance & Out of Pocket | 8 | 12% |  |
| Out of Pocket | 5 | 8% |  |
| Insurance & Medications Purchased Abroad | 1 | 2% |  |
| * Study data for % of total population of adults and children;<br>Comparison Adult US prevalence drawn from <sup>11-14</sup> |  |  |  |
| <sup>+</sup> Reported metrics for COPD (6.7%), Asthma (9.5%) § Migraine (15.3%) and ▼ Anemia (5.6%). |  |  |  |

Respondents reported 62 (21%) persons who required daily medication to manage chronic medical conditions and, of those, 17 (27%) reported difficulty acquiring needed medications. Most households, 51 (78.5%), reported that all household members used insurance to procure medications, while 9 households (13.8%) reported some household members using insurance while others paid for medications out-of-pocket, and 5 households (7.7%) reported that all household members paid for medication costs as out-of-pocket expense (Table 2).

Respondents reported a high level of health insurance coverage with 285 persons (97%) with some form of health insurance at least six months after the end of Refugee Medical Assistance (Table 1). Household members with health insurance were predominantly covered by Medicaid, with 55 out of 65 households (85%) relying on the CT Husky State Medicaid as their sole form of health insurance, and an additional 7 households relying on a mix of Medicaid and other forms of insurance such as private or employer-based plans. Only 1 household relied solely on an employer-based insurance and 2 households were uninsured.

Resettled households were integrated into the primary care system with 53 households (81.5%) that reported a primary care physician as their main method of obtaining health services. An additional 6 (9.2%) households primarily utilized episodic care in the form of urgent care and 5 households (7.7%) reported accessing the emergency department primarily for care. Households reported a median of 5 healthcare visits in the 6-months prior with 4 to the primary physician and one acute care visit (Table 2).

Resettled households reported a high median satisfaction care, albeit with very wide variation in each domain (Table 3). The PSQ-18 median scores by domain were General Satisfaction, 4.5 (1.5-5.0); Technical Quality, 4.0 (2.3-5.0); Interpersonal manner, 4.5 (2.5-5); Communication, 4.5 (2.0-5.0); Financial Aspects, 4.5 (1.0-5.0); Time spent with doctor, 4 (1.0-5.0); and Accessibility of services, 3.5 (1.8 – 5.0).

**Table 3:**
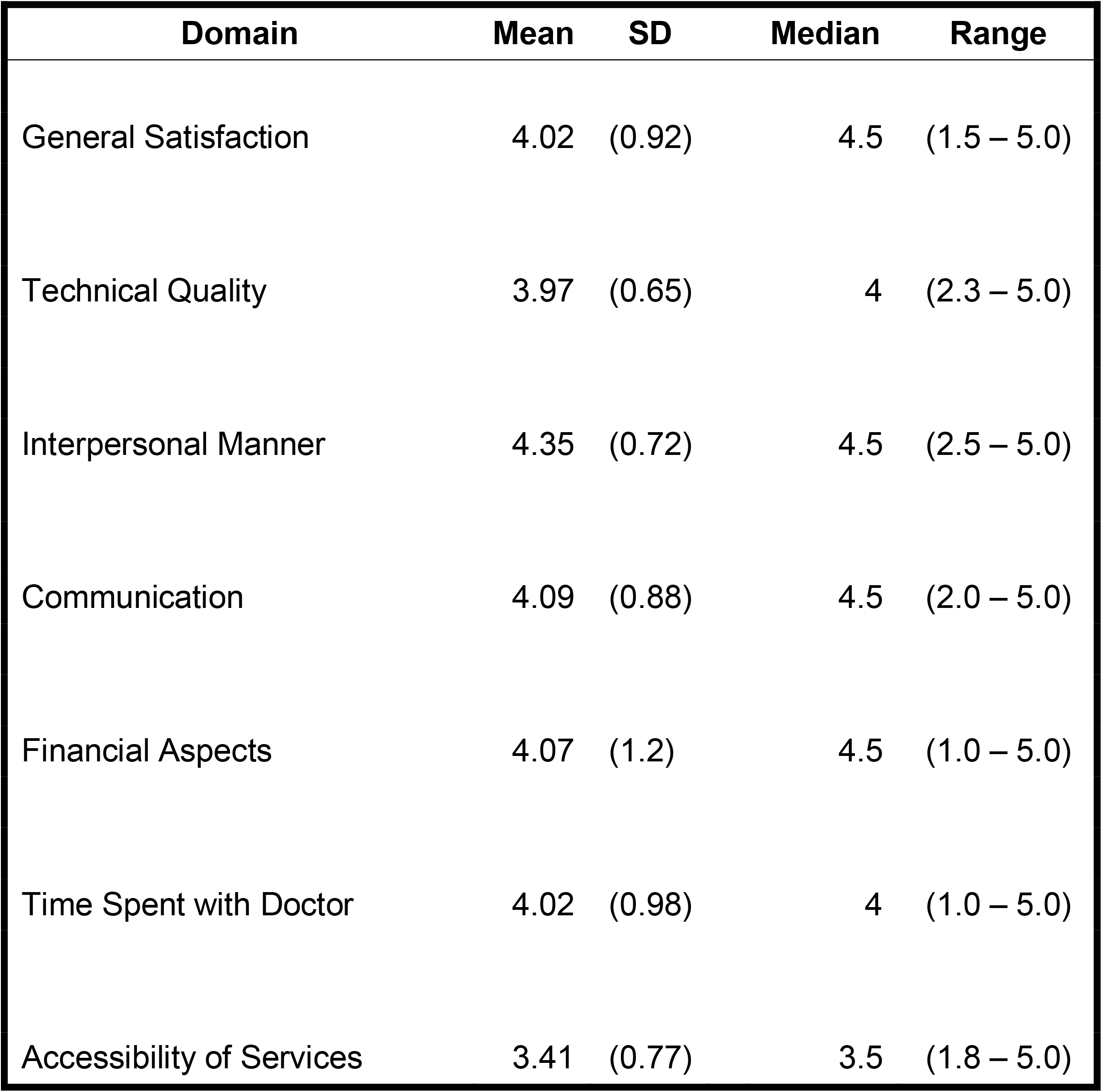
PSQ-18 Scores by Domain.

## DISCUSSION

The Arabic-speaking refugee population in this survey was young, with an age breakdown similar to that of all refugees arriving in the same time period,[18] but younger than that of the US population at large. Forty-eight percent of refugees represented in this study were under the age of 18, compared to 24% for the US population[19], and 61% were under the age of 25. Only 6 of the 295 (2%) Arabic-speaking refugees surveyed were over the age of 65.

Forty-seven households responded that at least one member of the household had a chronic medical condition (72%). Studies targeting adult refugee populations have documented similar rates of chronic non-communicable disease (NCD) as the US population, with up to 51% of refugee adults having at least once NCD, and 9.5% having three or more NCDs[20, 21]. Specifically, rates of hypertension, diabetes, and hyperlipidemia in refugee populations, at 24.1%, 7.8%, and 27.1%, respectively, have been noted to be comparable to the US population at large, however, these conditions were medically less controlled in the refugee population[22]. Despite similarities between refugee populations and the US population at large, variation in the prevalence and types of chronic conditions among different refugee groups have been noted, with one study demonstrating significant differences in prevalence of chronic disease based on location of origin[23]. The amount of time living in the USA prior to interview could also impact rates of chronic conditions, with increases in both obesity and hypertension noted in refugees with increased length of stay in the United States[24].

Given this documented high prevalence of chronic NCDs amongst refugee populations resettled in the USA, access to routine healthcare and daily medications to control chronic conditions is paramount to maintaining community health and reducing the cost of case management. In our study, Arabic-speaking refugee households were almost all covered by some form of medical insurance – with most households covered by State Medicaid. A similar result was found in the Iraqi refugee population in Michigan, where 100% of refugees surveyed one year after arrival were covered by Medicaid[25].

Conversely, 86.6% of refugees in San Antonio, Texas did not have any form of health insurance[26]. This variation in health coverage is likely attributable to differences in Medicaid eligibility requirements from state to state[27], with states adopting Medicaid expansion under the Affordable Care Act, such as Michigan and Connecticut, providing increased access to healthcare for refugees in comparison to states that have not adopted the expansion, such as Texas. Connecticut has relatively broad inclusion criteria for Medicaid eligibility that includes all children under the age of 18, caretakers of children, and low-income adults under the age of 65 that meet income thresholds,[28] contributing to a lower uninsured rate of 5% compared to the 9% uninsured rate for the United States[29]. The high rate of Medicaid coverage among refugees in states with Medicaid expansion may be impacted in the future by Trump administration guidelines that could prevent those who access public benefits from being eligible for permanent residence and citizenship – with the effect of discouraging refugees from submitting Medicaid applications. One projection estimates that millions of children will lose health coverage as a result of the rule change as new applications decrease and parents using existing services decide to unenroll to maintain eligibility for citizenship[30]. This could have profound effects on resettled refugees, especially in the era of COVID-19.

Respondents reported high levels of median satisfaction as measured by the PSQ-18. The very high levels of access to health insurance in our sample may have contributed to overall satisfaction as many barriers to access and payment for health services are mitigated by inclusion in the Medicaid program. While there were high *median* scores in each domain, there was wide variability in scores reported for each domain including several minimum scores. Our study was not powered to conduct sub-group analyses to identify the factors associated with low score reports in each domain. Additional work is needed in the form of qualitative interviews or focus groups to better understand the reasons underlying these negative perceptions of care received.

The domain with the lowest median score was accessibility of services. Similar findings were reported in another study that also used the PSQ-18 to survey Vietnamese refugees resettled in the US[31]. In that study, a majority of respondents had favorable views of their healthcare but indicated that language barriers made it challenging to access care. Several factors may contribute to these barriers in accessing care, including acculturation, lack of reliable transportation, difficulty navigating the complex US healthcare system, and language and communication barriers[3-5, 32, 33].

The difficulty acquiring prescription medications for refugees in this study despite widespread insurance coverage warrants further study. A recent systematic review on access to prescription medication and pharmacy services among resettled refugees in Australia found that while there was a paucity of research in this area, a wide variety of factors including language and cultural barriers, difficulty navigating the system for obtaining prescription medications (e.g. may be used to simply purchasing directly from a pharmacist), use of traditional medicine and medication non-adherence all contribute to decreased access to medications[34].

### Limitations

This study has several important limitations that must be considered. First, while every attempt was made to contact resettled persons, 27 of the original 117 households obtained from IRIS had no contact information or were known to have returned to their home countries. We had no baseline information on these households and cannot assess how similar they are to the respondent households. It may be that these households who are disconnected from the refugee resettlement agency are those that are having greater difficulty obtaining health insurance and accessing health services.

Further, while we report a lower prevalence of chronic condition in the *overall* respondent population the survey instrument did not clearly request the number of *adults* with chronic conditions. While the authors believe that this was the common understanding amongst respondents, the lack of precision on this point leads to a lower reported percentage. The percentage of households reporting a household member with a chronic condition (72%) is substantially higher than the host community. As such, if the number of persons with chronic conditions are assumed to all be adults, then the numbers more closely mirror or exceed the rates in the host community (hypertension – 12.3% vs 27.6%; Diabetes – 11.0% vs 8.4%; chronic respiratory conditions – 13.6% vs COPD 6.2% or asthma 9.5%; hematologic disorders – 4.5% vs 2%).

Furthermore, refugee respondents were aware that the interviewers were part of the health system where they receive care and that their contact information was provided by IRIS, where they receive social support. In this context, respondents may have provided more positive perceptions of their care in deference to this known connection.

Lastly, the degree of health insurance coverage in our study may limit the ability to generalize the perceptions of healthcare to other settings with poorer access to health insurance for resettled persons. As previously mentioned, most refugee respondents in this study were covered by Medicaid, the eligibility requirements and quality of which vary from state to state[27]. Roughly 40% of refugees are resettled to states that have not adopted Medicaid expansion under the ACA[35]. Studying PSQ-18 results among Arabic-speaking refugees with no insurance in these states may help elucidate how geography of landing impacts perceptions and delivery of healthcare for refugees.

## CONCLUSION

Arabic-speaking persons resettled in CT are young and have adult rates of chronic medical conditions similar to the host community. A high percentage of resettled households reported coverage by medical insurance and the majority reported state Medicaid as their primary insurance. Respondents expressed median high satisfaction with health services but with wide variation. Accessibility of health services in Arabic and ability to obtain medications remain areas in need of improvement for resettled refugees in CT. Additional research is needed to understand the factors that contribute to poor perceptions of health services in patients with this high access to health insurance. Furthermore, additional comparative research is needed to assess the impact of access to state-based health insurance on access to and perceptions of healthcare services for resettled persons.

## Data Availability

Data available upon request of the authors

## ACKNOWLEDGEMENTS

The authors are grateful for the support of the Program on Refugees, Forced Displacement, and Humanitarian Responses and the Whitney and Betty MacMillan Center for International and Area Studies at Yale University in funding this study. The authors would further like to express their thanks and appreciation to Leslie Koons and her team at Integrated Refugee and Immigrant Services, CT for their assistance in this project.

