## Supplemental File 1 for "Utilization, satisfaction and barriers to access of health services for Arabic-speaking refugees resettled in Connecticut, USA after completion of Refugee Medical Assistance (RMA)"

Appendix – Survey Instrument

Start of Block: General Questionnaire

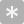

Q1 Has consent been obtained?

- Yes (1)
- No (2)

| Page Break |
| --- |

Q32 Person conducting survey

- Susan Aboeid (1)
- Sarah Aly (2)
- Salah Mankash (3)
- Aminah Sallam (4)
- Ali Elreichouni (5)
- Islam Salem (7)
- Noah Ghossein (9)

Q1 Study ID

________________________________________________________________

| Page Break |
| --- |

Q2 Number of Members of Household

Adults : _______ (1)

Children : _______ (2)

Total : ________

| Page Break |
| --- |

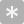

Q3 Ages of Household Members

Under 18 : _______ (1)

18 - 24 : _______ (2)

25 - 34 : _______ (3)

35 - 44 : _______ (4)

45 - 54 : _______ (5)

55 - 64 : _______ (6)

65 - 74 : _______ (7)

75 - 84 : _______ (8)

85 or older : _______ (9)

Total : ________

| Page Break |
| --- |

Q4 How many members of the household have health insurance?

________________________________________________________________

Q5 What type of medical insurance does your family have (select all that apply)?

- Medicaid/Husky/State health insurance (1)
- Insurance provided by work (2)
- Private insurance purchased directly by family (3)
- No insurance (4)

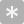

Q6 How many members of your household have been diagnosed with the following conditions?

High blood pressure : _______ (1)

Diabetes : _______ (2)

Heart problems : _______ (3)

Problems breathing or Chronic respiratory problems : _______ (4)

High cholesterol : _______ (5)

Chronic headaches : _______ (6)

Blood related conditions (Hematologic) : _______ (7)

Cancer : _______ (8)

Other chronic conditions : _______ (9)

Total : ________

| Page Break |
| --- |

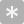

Q7 How many members of the household have difficulty walking or performing normal daily activities?

________________________________________________________________

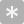

Q8 How many members of the household are supposed to take medications on a daily basis (even if they don't take it every day)?

________________________________________________________________

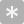

Q9 How many members of the household have difficulty getting the daily medications they need?

________________________________________________________________

| Page Break |
| --- |

Q10 How do household members usually pay for medications (select all that apply)?

- Cash/Out of pocket (1)
- Insurance pays for all or part of medicine costs (family pays a co-payment) (2)
- Free samples from a doctor's office (3)
- Medications purchased out of the country by friend or family (4)
- Other (5)

Q11 What is the primary method that members of the family seek medical care?

- Primary doctor (1)
- Walk-in clinic/Urgent care (2)
- Emergency department (3)
- Other (4)

Q12 In the last 6 months, how many times has a member of the household visited one of the following?

Primary doctor : _______ (1)

Walk-in clinic/Urgent care : _______ (2)

Emergency department : _______ (3)

Other : _______ (4)

Total : ________

End of Block: General Questionnaire

Start of Block: Patient Satisfaction Questionnaire (PSQ-18)

Q13 How strongly do you agree or disagree with the following statement?

|  | Strongly Agree (1) | Agree (2) | Neither agree nor disagree (3) | Disagree (4) | Strongly Disagree (5) |
| --- | --- | --- | --- | --- | --- |
| Doctors are good about explaining the reasons for medical tests (1) |  |  |  |  |  |

Q14 How strongly do you agree or disagree with the following statement?

|  | Strongly agree (1) | Agree (2) | Neither agree nor disagree (3) | Disagree (4) | Strongly disagree (5) |
| --- | --- | --- | --- | --- | --- |
| My doctors office has everything needed to provide complete medical care (1) |  |  |  |  |  |

Q15 How strongly do you agree or disagree with the following statement?

|  | Strongly agree (1) | Agree (2) | Neither agree nor disagree (3) | Disagree (4) | Strongly disagree (5) |
| --- | --- | --- | --- | --- | --- |
| The medical care I have received has been almost perfect (1) |  |  |  |  |  |

Q16 How strongly do you agree or disagree with the following statement?

|  | Strongly Agree (1) | Agree (2) | Neither agree nor disagree (3) | Disagree (4) | Strongly disagree (5) |
| --- | --- | --- | --- | --- | --- |
| Sometimes I wonder if my diagnosis is correct (1) |  |  |  |  |  |

Q17 How strongly do you agree or disagree with the following statement?

|  | Strongly agree (1) | Agree (2) | Neither agree nor disagree (3) | Disagree (4) | Strongly disagree (5) |
| --- | --- | --- | --- | --- | --- |
| I feel confident that I can get medical care when I need it without financial difficulty (1) |  |  |  |  |  |

Q18 How strongly do you agree or disagree with the following statement?

|  | Strongly agree (1) | Agree (2) | Neither agree nor disagree (3) | Disagree (4) | Strongly disagree (5) |
| --- | --- | --- | --- | --- | --- |
| When I go for medical care the doctors are careful to check everything when treating and examining me (1) |  |  |  |  |  |

Q19 How strongly do you agree or disagree with the following statement?

|  | Strongly agree (1) | Agree (2) | Neither agree nor disagree (3) | Disagree (4) | Strongly disagree (5) |
| --- | --- | --- | --- | --- | --- |
| I have to pay more than I can afford for my medical care (1) |  |  |  |  |  |

Q20 How strongly do you agree or disagree with the following statement?

|  | Strongly agree (1) | Agree (2) | Neither agree nor disagree (3) | Disagree (4) | Strongly disagree (5) |
| --- | --- | --- | --- | --- | --- |
| I have easy access to medical specialists when I need them (1) |  |  |  |  |  |

Q21 How strongly do you agree or disagree with the following statement?

|  | Strongly agree (1) | Agree (2) | Neither agree nor disagree (3) | Disagree (4) | Strongly disagree (5) |
| --- | --- | --- | --- | --- | --- |
| Where I get medical care, people have to wait too long for emergency treatment (1) |  |  |  |  |  |

Q22 How strongly do you agree or disagree with the following statement?

|  | Strongly agree (1) | Agree (2) | Neither agree nor disagree (3) | Disagree (4) | Strongly disagree (5) |
| --- | --- | --- | --- | --- | --- |
| Doctors are too impersonal or business-like towards me (1) |  |  |  |  |  |

Q23 How strongly do you agree or disagree with the following statement?

|  | Strongly agree (1) | Agree (2) | Neither agree nor disagree (3) | Disagree (4) | Strongly disagree (5) |
| --- | --- | --- | --- | --- | --- |
| My doctors treat me in a very friendly and courteous manner (1) |  |  |  |  |  |

Q24 How strongly do you agree or disagree with the following statement?

|  | Strongly agree (1) | Agree (2) | Neither agree nor disagree (3) | Disagree (4) | Strongly disagree (5) |
| --- | --- | --- | --- | --- | --- |
| Sometimes I feel that my doctors hurry too much when they treat me (1) |  |  |  |  |  |

Q25 How strongly do you agree or disagree with the following statement?

|  | Strongly agree (1) | Agree (2) | Neither agree nor disagree (3) | Disagree (4) | Strongly disagree (5) |
| --- | --- | --- | --- | --- | --- |
| Doctors sometimes ignore what I tell them (1) |  |  |  |  |  |

Q26 How strongly do you agree or disagree with the following statement?

|  | Strongly agree (1) | Agree (2) | Neither agree nor disagree (3) | Disagree (4) | Strongly disagree (5) |
| --- | --- | --- | --- | --- | --- |
| I have some doubts about the abilities of the doctors that treat me (1) |  |  |  |  |  |

Q27 How strongly do you agree or disagree with the following statement?

|  | Strongly agree (1) | Agree (2) | Neither agree nor disagree (3) | Disagree (4) | Strongly disagree (5) |
| --- | --- | --- | --- | --- | --- |
| Doctors usually spend enough time with me at my visits (1) |  |  |  |  |  |

Q28 How strongly do you agree or disagree with the following statement?

|  | Strongly agree (1) | Agree (2) | Neither agree nor disagree (3) | Disagree (4) | Strongly disagree (5) |
| --- | --- | --- | --- | --- | --- |
| I find it hard to get an appointment for medical care right away (1) |  |  |  |  |  |

Q29 How strongly do you agree or disagree with the following statement?

|  | Strongly agree (1) | Agree (2) | Neither agree nor disagree (3) | Disagree (4) | Strongly disagree (5) |
| --- | --- | --- | --- | --- | --- |
| I am dissatisfied with some things about the medical care I receive (1) |  |  |  |  |  |

Q30 How strongly do you agree or disagree with the following statement?

|  | Strongly agree (1) | Agree (2) | Neither agree nor disagree (3) | Disagree (4) | Strongly disagree (5) |
| --- | --- | --- | --- | --- | --- |
| I am able to get medical care whenever I need it (1) |  |  |  |  |  |

End of Block: Patient Satisfaction Questionnaire (PSQ-18)

Start of Block: Gift Card Receipt

Q33 Was the gift card given in person?

- Yes (1)
- No (2)

Display This Question:

If Was the gift card given in person? = No

Q34 Please enter the address where you'd like to receive your gift card

________________________________________________________________

________________________________________________________________

________________________________________________________________

________________________________________________________________

________________________________________________________________

Display This Question:

If Was the gift card given in person? = Yes

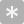

Q35 Please enter the last 4 digits of the gift card

________________________________________________________________

End of Block: Gift Card Receipt
